# Trends and variation in unsafe prescribing of methotrexate: a cohort study in English NHS primary care

**DOI:** 10.1101/19000919

**Authors:** Brian MacKenna, Helen J Curtis, Alex J Walker, Richard Croker, Seb Bacon, Ben Goldacre

## Abstract

**Objective:** To describe trends and geographical variation in methotrexate prescribing that breaches national safety recommendations; deaths from methotrexate poisoning; and associated litigation.

**Methods:** A retrospective cohort study of English NHS primary care prescribing data, complemented by information obtained through Freedom of Information (FOI) requests. The main outcome measures were: (1) variation in ratio of breaching / adherent prescribing, geographically and over time, between General Practices and Clinical Commissioning Groups; (2) description of responses to FOI requests.

**Results:** Out of 7349 NHS General Practices in England, 1689 practices prescribed both 2.5mg and 10mg tablets to individual patients in 2017, breaching national guidance. In April 2018, 697 practices (at the 90th centile and above) prescribed at least 14.3% of all methotrexate as 10mg tablets, breaching national guidance. The 66 practices at the 99th percentile and above gave at least 52.4% of all prescribed methotrexate in the form of 10 mg tablets. The prescribing of 10mg tablets has fallen over 7 years, with 10mg tablets as a proportion of all methotrexate tablets falling from 9.1% to 3.4%. 21 deaths caused by methotrexate poisoning have been reported from 1993-2017.

**Conclusions:** The prevalence of unsafe methotrexate prescribing has reduced but it remains common, with substantial variation between organisations. We recommend the NHS invests in better strategies around implementation of safety recommendations. 21 deaths have been attributed to methotrexate poisoning but with no further details easily available: the full coroners reports for these deaths should be reviewed to identify recurring themes.

## Introduction

Methotrexate is a folic acid antagonist, commonly prescribed for conditions such as rheumatoid arthritis, Crohn’s disease, severe psoriasis and some cancers. Methotrexate has a narrow therapeutic index, and higher doses can result in potentially fatal adverse events including blood disorders, liver toxicity, and shortness of breath.[1] Due to its narrow therapeutic index, and an unusual once-weekly dosing regimen, this drug presents a particularly high risk of accidental overdose: patients may incorrectly take methotrexate on a daily basis; and overdose may occur due to dispensing or prescribing errors. The British National Formulary (BNF) states that methotrexate should only usually be prescribed in a single strength of tablet, usually 2.5mg, to reduce the risk of harm from errors.

An NHS inquiry was convened in 2000 following the death of a patient who had accidentally taken a high dose of methotrexate daily. The inquiry made 28 recommendations to minimise the future risk of harm [2] and the (now defunct) National Patient Safety Agency (NPSA - see box 1) issued a national level patient safety alert in 2006 accompanied by a series of guides and recommendations to reduce the risk of harm to patients.[3] Similar incidents have occurred across Europe and due to the persistence of such issues, the European Medicines Agency (EMA) has launched a review of methotrexate dosing errors.[4]

### Box 1: Explainer: Organisations who are featured in this article

#### Clinical Commissioning Group (CCG)

A collection of GP practices working together to plan, commission, and pay for health care services in a local area. There are approximately 200 CCGs in England.

#### European Medicines Agency (EMA)

An agency of the Europeaan Union responsible for evaluation of medicines. They monitor and supervise the safety of medicines that have been authorised in the EU, to ensure that their benefits outweigh their risks.

#### Office of National Statistics (ONS)

the UK’s largest independent producer of official statistics.

#### NHS Resolution

an NHS body advising the NHS on how to resolve compensation claims fairly, while sharing insights for quality improvement, and preserving resources for patient care.

#### NHS Business Services Authority

an NHS body which calculates the remuneration and reimbursement due to pharmacies across England, and publishes data on prescribing.

#### NHS Improvement

an NHS body with financial regulatory responsibility for NHS trusts and independent providers of NHS-funded care. It also has responsibility for patient safety across the NHS.

#### National Patient Safety Agency (NPSA)

The NPSA was an arm’s length body of the Department of Health with a mandate to identify patient safety issues and find appropriate solutions. It was abolished in 2012 with patient safety issues transferred to NHS England, and then to NHS Improvement in April 2016.

Correct prescribing of methotrexate remains a key priority for the NHS. It is one of only 16 targeted issues in the current NHS “Never Events” list (Box 1).[5] In addition, NHS Improvement have issued further guidance on Never Events to supplement the work of the NPSA: here the correct prescribing of methotrexate is one of only 11 targeted issues.[6] The NHS Improvement document states that: “All electronic prescribing and dispensing software programmes in primary and secondary care locations must include oral methotrexate alerts and prompts.” Surprisingly, however, there is no mention of monitoring compliance in routine data, or feeding back to practices and Clinical Commissioning Groups (CCGs - see box 1) if a breach of guidance is identified.

Our OpenPrescribing.net service is a publicly funded and openly accessible explorer for NHS primary care prescribing data, launched in 2015, with 100,000 unique users in the past year including doctors, pharmacists and patients. It supports complex bespoke data queries, and displays numerous predefined standard measures for safety, cost, and effectiveness for every practice in England. OpenPrescribing has a standard measure for methotrexate prescribing [7] showing the proportion prescribed in potentially dangerous 10mg tablet doses. We have noted a substantial number of practices and CCGs are commonly in breach of best practice guidance around methotrexate.

We therefore set out to: describe the long-term trends in methotrexate prescribing over time; describe variation between practices and CCGs in their implementation of the safety guidance; describe and map current variation at CCG and practice level; and describe the harm associated with methotrexate errors at a national level.

## Methods

### Study design

We analysed prescribing practice by conducting a retrospective cohort study in prescribing data from all English NHS general practices and CCGs. We assessed harm associated with methotrexate by requesting information through Freedom of Information (FOI) requests to the Office for National Statistics (ONS - see box 1) and NHS Resolution (see - box 1).

### Data Sources

We extracted data from the OpenPrescribing.net database. This imports openly accessible prescribing data from the large monthly files published by the NHS Business Services Authority (see box 1) which contain data on cost and items prescribed for each month, for every typical general practice and CCG in England since mid-2010.[8] The monthly prescribing datasets contain one row for each different medication and dose, in each prescribing organisation in NHS primary care in England, describing the number of items (i.e. prescriptions issued) and the total cost. These data are sourced from community pharmacy claims data and therefore contain all items that were dispensed. We extracted all available prescribing data for institutions identified as general practices, and excluded all other organisations such as prisons. The number of patients registered at each practice was obtained from NHS Digital data.[9]

In addition we requested data covered by FOI [10] to the NHS Business Services Authority for aggregated patient level data in order to ascertain where co-prescribing of both methotrexate 2.5mg and 10mg tablets occurred for an individual patient.

We also sent access requests for data covered by FOI to ONS and NHS Resolution. Briefly we asked for data relating to deaths from methotrexate (ONS) [11] and associated legal claims and costs (NHS Resolution). These requests and responses can be viewed on Figshare.[12]

### Methotrexate Prescribing

We extracted data on all prescriptions dispensed between August 2010 and April 2018, the latest data available when we started our analysis, for prescribing of methotrexate of any form, using BNF codes starting with 0801030P (injections, ampoules and pre-filled pens) and 1001030U0 (tablets, liquids and pre-filled pens). We excluded liquids due to the low volume of prescribing. We calculated CCG and practice-level deciles at each month for the proportion of total methotrexate tablets prescribed as 10mg tablets and plotted them on a time series chart. Additionally we analysed data supplied from the NHS Business Services Authority on prescribing of both 2.5mg and 10mg tablets to individual patients.

### Geographical variation at CCG level across England

We created choropleth maps of the overall proportion of methotrexate tablets prescribed as 10mg tablets between May 2017 and April 2018 for each CCG in England.

### Factors associated with prescribing of methotrexate 10mg tablets

We examined factors associated with a high proportion (>10%) of 10mg methotrexate tablet prescribing, using a mixed effects logistic regression model. We selected variables from data available on individual CCGs and practices from publicly available data that have previously been shown to be associated with variation in prescribing. These variables were: Index of Multiple Deprivation; Quality Outcomes Framework Score; a composite prescribing score (determined by taking the mean percentile of the current OpenPrescribing measures-see appendix);[12] the primary electronic health record (EHR) system used in the practice; whether a practice was single-handed; the urban/ruralness of the practice; proportion of patients over 65; proportion of patients under 18; and the proportion of patients with a long term health condition. We explored the impact of CCG as a random effect to estimate the influence of CCG membership on individual practices within their organisation. Continuous variables were categorised a priori into quintiles in order to allow for nonlinearity of effects and to enhance the intelligibility of results.

The outcome used was a binary variable of whether a practice had >10% of methotrexate prescriptions issued with 10mg tablets. We selected this threshold a priori since the majority of practices had no 10mg methotrexate prescribing, but we did not want to include practices that only very occasionally prescribe 10mg doses. The model was used to calculate odds ratios and 95% confidence intervals (CI) for each of the fixed effect variables, as well as an R-squared value (along with the significance level) to describe the degree of variance associated with CCG membership.

### Harms associated with methotrexate errors at a national level

Responses to our FOI requests were aggregated and summarised.

### Software and Reproducibility

Data management was performed using Python 3 and Google BigQuery, with analysis carried out using Stata 14.2 and Python. Data, as well as all code for data management and analysis, is archived online and available for free re-use, including as a Jupyter notebook.[13]

### Patient and Public Involvement

Our website OpenPrescribing.net, is an openly accessible data explorer for all NHS England primary care prescribing data, which receives a large volume of user feedback from professionals, patients and the public. This feedback is used to refine and prioritise our informatics tools and research activities. Patients were not formally involved in developing this specific study design.

## Results

### Methotrexate prescribing

Methotrexate 10mg tablets represent 3.4% of all methotrexate tablet prescribing; this has reduced from 9.1% since October 2010. Figure 1 shows the trends and variation in prescribing of 10mg tablets as a proportion of all methotrexate tablets across England’s practices and CCGs between 2010 and 2018. Although the general trend is downwards, there is still very extensive variation. In April 2018 most practices prescribed no methotrexate 10mg tablets (median 0%). However 697 NHS GP practices in England (at the 90th centile and above) prescribed at least 14.3% of all methotrexate as 10mg tablets, in breach of BNF guidance. The 66 practices at the 99th percentile and above gave at least 52.4% of all prescribed methotrexate in the form of 10 mg tablets.

**Figure 1.**
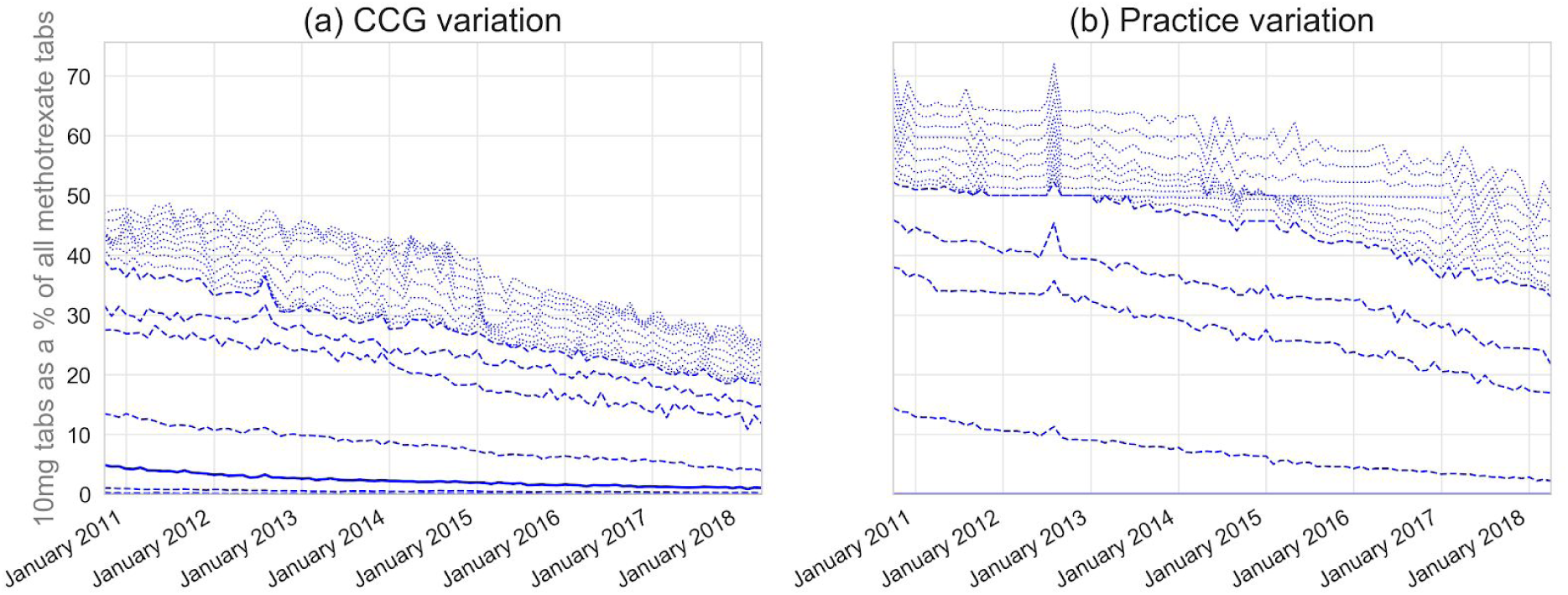
Prescribing of 10mg tablets as a proportion of all methotrexate tablets prescribed across CCGs (a) and practices (b) in England, October 2010 -April 2018

### Median is a solid line, dashed lines represent deciles, light dotted lines represent extreme percentiles (1-9, 90-99)

We further investigated co-prescribing of methotrexate 2.5mg and 10mg tablets in individual patients by submitting an FOI request to NHS Business Services Authority for the number of individual patients receiving both 10mg and 2.5mg tablets in the same prescription. For information governance reasons numbers of patients from 1-4 were suppressed at source. 1689 (23%, n=7349) practices co-prescribed both methotrexate 2.5mg and 10mg tablets to individual patients, against current safety guidance. 197 practices prescribed mixed strengths for more than 5 of their individual patients, affecting 1826 people.

### Geographical variation at CCG level across England

Figure 2 displays the variation in prescribing of 10mg tablets as a proportion of all tablets over 12 months across England (Mean 4%, Range 0% - 38%) and London (Mean 10%, Range 1%-29%). The ten CCGs with highest prescribing of 10mg methotrexate in breach of current BNF safety guidance are listed in Table 1, with the proportion of prescriptions that breached guidance.

**Table 1.** 10mg items as a proportion of all methotrexate tablets - Highest 10 CCGs May 2017 - April 2018

| CCG Name | (%) |
| --- | --- |
| NHS DARTFORD, GRAVESHAM AND SWANLEY CCG | 37.6 |
| NHS MILTON KEYNES CCG | 29.8 |
| NHS BEXLEY CCG | 28.6 |
| NHS ENFIELD CCG | 26.2 |
| NHS LEEDS CCG | 25.6 |
| NHS WALTHAM FOREST CCG | 21.4 |
| NHS GREENWICH CCG | 20.8 |
| NHS CENTRAL LONDON (WESTMINSTER) CCG | 17.9 |
| NHS HOUNSLOW CCG | 17.7 |
| NHS HILLINGDON CCG | 17.3 |

**Figure 2.**
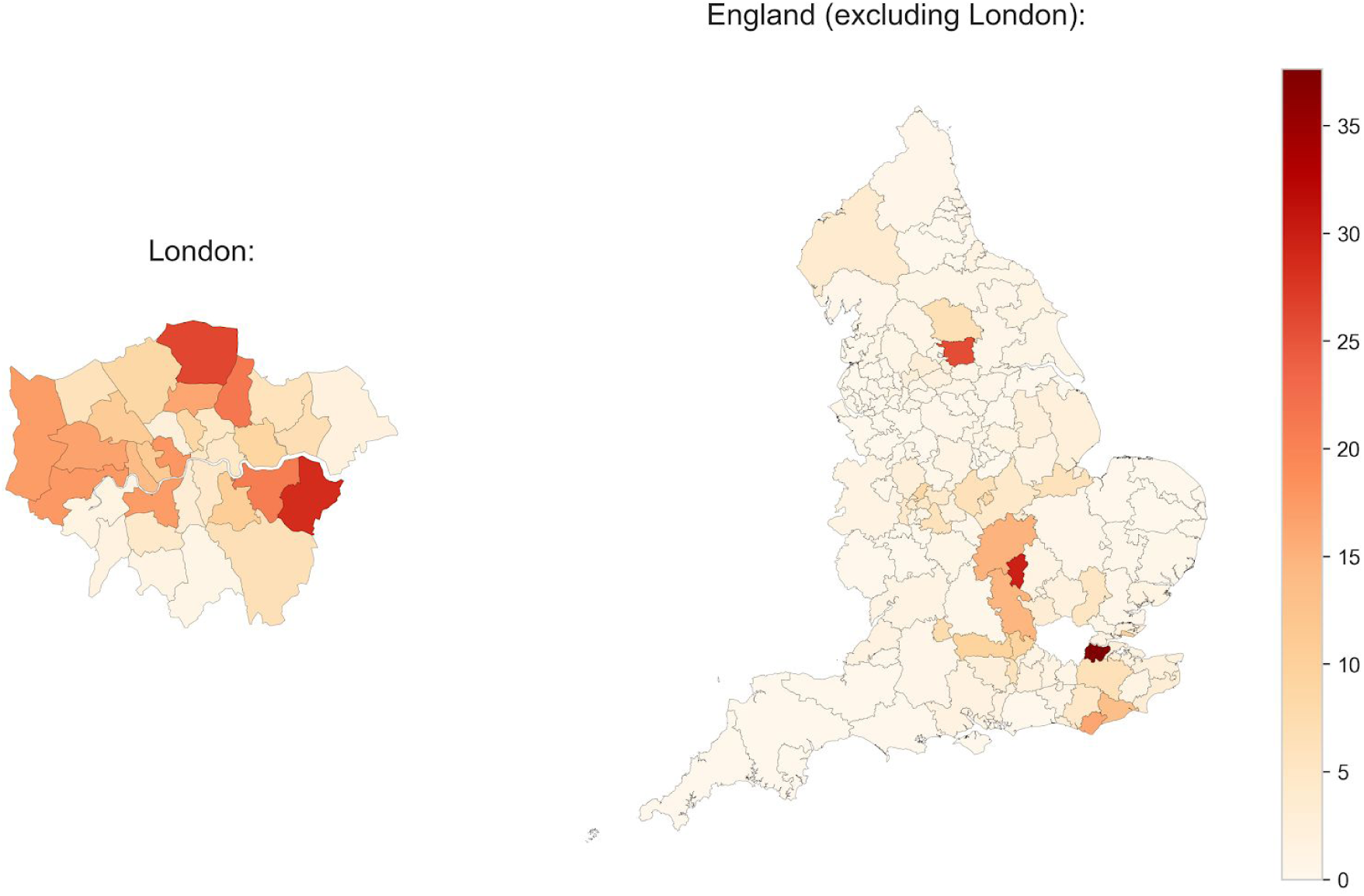
10mg items as a proportion of all methotrexate tablets - variation between CCGs May 2017 - April 2018

### Factors associated with prescribing of methotrexate 10mg

We modelled the practice factors associated with prescribing a high proportion (>10%) of methotrexate 10mg tablets (Table 2). Demographic factors were associated with prescribing methotrexate 10mg tablets: practices with a higher proportion of patients over 65 were less likely to have high 10mg prescribing (multivariable odds ratio for highest vs lowest: 0.27, 95% CI 0.16-0.45). Practices with a higher proportion of patients under 18 were also less likely to have high 10mg prescribing (odds ratio for highest vs lowest: 0.68, 95% CI 0.48-0.96). Urban areas were especially likely to have high 10mg tablet prescribing, but this is likely mostly due to a focus of 10mg tablet prescribing in London and Leeds (Figure 2).

**Table 2.** Absolute proportion of 10mg methotrexate prescribing, stratified by various practice factors, along with odds ratios from a univariable and multivariable logistic regression model. IMD, Index of Multiple Deprivation; QOF, Quality Outcomes Framework.

| Variable | Category boundaries | Proportion of practices with >10% 10mg methotrexate prescribing | Univariable logistic regression |  |  | Multivariable logistic regression |  |  |
| --- | --- | --- | --- | --- | --- | --- | --- | --- |
|  |  |  | Odds ratio | 95% CI |  | Odds ratio | 95% CI |  |
| % of patients over 65 | 0-10.9 | 27.2 | Ref |  |  | Ref |  |  |
|  | 10.9-15.5 | 19.3 | 0.64 | 0.54 | 0.76 | 0.76 | 0.59 | 0.99 |
|  | 15.5-18.9 | 13.6 | 0.42 | 0.35 | 0.51 | 0.63 | 0.45 | 0.89 |
|  | 18.9-22.5 | 8.1 | 0.23 | 0.19 | 0.29 | 0.36 | 0.24 | 0.55 |
|  | 22.5-92.2 | 5.4 | 0.15 | 0.12 | 0.20 | 0.27 | 0.16 | 0.45 |
| % of patients under 18 | 0-17.8 | 11.6 | Ref |  |  | Ref |  |  |
|  | 17.8-19.6 | 10.7 | 0.92 | 0.73 | 1.16 | 0.85 | 0.62 | 1.17 |
|  | 19.6-21.2 | 12.9 | 1.13 | 0.90 | 1.41 | 0.69 | 0.49 | 0.95 |
|  | 21.2-23.6 | 17.6 | 1.63 | 1.32 | 2.01 | 0.71 | 0.51 | 0.99 |
|  | 23.6-53.6 | 20.8 | 2.01 | 1.63 | 2.46 | 0.68 | 0.48 | 0.96 |
| % with a long term health condition | 16.5-47.0 | 23.6 | Ref |  |  | Ref |  |  |
|  | 47.0-51.5 | 18.4 | 0.73 | 0.61 | 0.87 | 0.88 | 0.69 | 1.12 |
|  | 51.5-55.3 | 12.7 | 0.47 | 0.39 | 0.58 | 0.87 | 0.66 | 1.15 |
|  | 55.3-59.7 | 10.6 | 0.38 | 0.31 | 0.47 | 0.81 | 0.60 | 1.09 |
|  | 59.7-96.0 | 8.3 | 0.29 | 0.24 | 0.37 | 0.78 | 0.56 | 1.09 |
| Single handed practice | No | 14.6 | Ref |  |  | Ref |  |  |
|  | Yes | 15.9 | 1.11 | 0.87 | 1.42 | 1.21 | 0.87 | 1.70 |
| Urban/rural | Urban with major conurbation | 24.1 | Ref |  |  | Ref |  |  |
|  | Urban with minor conurbation | 1.3 | 0.04 | 0.02 | 0.11 | 0.12 | 0.02 | 0.62 |
|  | Urban with city and town | 10.2 | 0.36 | 0.30 | 0.42 | 0.18 | 0.10 | 0.35 |
|  | Urban with significant rural | 9.3 | 0.32 | 0.25 | 0.41 | 0.13 | 0.06 | 0.26 |
|  | Largely rural | 5.3 | 0.18 | 0.13 | 0.25 | 0.15 | 0.07 | 0.31 |
|  | Mainly rural | 5.0 | 0.17 | 0.11 | 0.24 | 0.15 | 0.07 | 0.33 |
| IMD | Least deprived | 12.0 | Ref |  |  | Ref |  |  |
|  |  | 12.5 | 1.05 | 0.84 | 1.31 | 1.04 | 0.75 | 1.44 |
|  |  | 16.5 | 1.46 | 1.18 | 1.80 | 0.98 | 0.69 | 1.38 |
|  |  | 16.9 | 1.49 | 1.21 | 1.84 | 0.99 | 0.67 | 1.46 |
|  | Most deprived | 15.7 | 1.37 | 1.11 | 1.69 | 1.19 | 0.77 | 1.84 |
| QOF | 14-523 | 19.0 | Ref |  |  | Ref |  |  |
|  | 523-541 | 17.3 | 0.89 | 0.74 | 1.07 | 0.89 | 0.69 | 1.14 |
|  | 541-550 | 13.8 | 0.68 | 0.56 | 0.83 | 0.77 | 0.59 | 1.00 |
|  | 550-557 | 13.1 | 0.64 | 0.52 | 0.78 | 0.80 | 0.61 | 1.04 |
|  | 557-559 | 10.0 | 0.47 | 0.38 | 0.58 | 0.65 | 0.48 | 0.88 |
| Composite prescribing score (lower is better) | <38.7 | 15.3 | Ref |  |  | Ref |  |  |
|  | 38.7-43.5 | 14.5 | 0.94 | 0.76 | 1.15 | 1.26 | 0.96 | 1.66 |
|  | 43.5-47.8 | 13.4 | 0.86 | 0.70 | 1.05 | 1.39 | 1.04 | 1.87 |
|  | 47.8-52.4 | 13.8 | 0.89 | 0.72 | 1.09 | 1.96 | 1.44 | 2.67 |
|  | >52.4 | 16.4 | 1.09 | 0.89 | 1.33 | 3.28 | 2.35 | 4.57 |
| Computer system | EMIS | 14.1 | Ref |  |  | Ref |  |  |
|  | Evolution | 1.7 | 0.11 | 0.01 | 0.78 | 1.57 | 0.15 | 16.58 |
|  | SystmOne | 13.9 | 0.98 | 0.85 | 1.13 | 0.97 | 0.70 | 1.34 |
|  | Vision | 29.5 | 2.56 | 2.03 | 3.24 | 1.68 | 1.07 | 2.65 |

Having a higher (worse) composite OpenPrescribing score was associated with a greater likelihood of high 10mg tablet prescribing. Prescribing practice largely did not correlate with the principal EHR system that a practice uses, with the exception of Vision practices, which were more likely to have high 10mg tablet prescribing (odds ratio for Vision vs EMIS: 1.68, 95% CI 1.07-2.65). The CCG to which a practice belongs (as a random effect) was significantly associated with high-dose prescribing (p <0.0001) and accounted for 25.5% of the variation in methotrexate 10mg tablet prescribing.

### Harms associated with methotrexate errors at a national level

In their FOI response, ONS data showed 21 reported deaths from 1993-2017 classified as a poisoning where methotrexate was the only drug mentioned on the death certificate. In total there were 24 deaths due to poisoning where methotrexate was mentioned on death certificates in England and Wales. The data reported by ONS and NHS Resolution was not sufficient to explore in any detail how methotrexate was involved in the reported death.[14]

## Discussion

### Summary

At least 21 people have died from methotrexate poisoning in England and Wales since 1993. Methotrexate 10mg tablets remain common but practice is extremely variable: most practices prescribed none; but 697 NHS GP practices in England prescribed at least 14.3% of all methotrexate as 10mg tablets; and 66 practices gave at least 52.4%. Breaches were more common in urban practices, and practices with a worse composite prescribing quality score. CCG membership explained 25.5% of prescribing variation, suggesting that CCGs exert a substantial influence on clinical practice around methotrexate.

### Strengths and weaknesses

Our prescribing data includes all prescribing in all typical practices in England, thus minimising the potential for obtaining a biased sample. We used real prescribing and spending data which are sourced from pharmacy claims and therefore did not need to rely on surrogate measures. This was complemented with aggregated patient level data to identify patterns of co-prescribing, which again was sourced from pharmacy claims. Using primary data, rather than survey data, eliminates the possibility of recall bias. We would ideally have included hospital prescribing, and have advocated for this data to be more widely shared: however at present it is only available for pharmaceutical industry marketing, and a limited range of unpublished analyses at NHS Improvement.

Data on deaths related to methotrexate was obtained from the ONS, the most robust dataset available on drug poisonings. This dataset is comprehensive but suffers from delays: figures are for deaths registered in each calendar year, rather than occurring in each year; and a coroner’s inquest can take months or even years to complete. The data on legal claims obtained from NHS Resolution has limited structure and does not give any information on the reason for the claim: deaths data may include deaths not related to primary care prescribing, e.g. after prescriptions that were given in hospital, or adverse reactions not caused by excessive dosing.

### Findings in Context

We are aware of no prior work on the prevalence of breaches on methotrexate guidance: however incomplete implementation of this important national NHS safety alert is consistent with extensive prior work showing incomplete or slow adoption for other national prescribing guidance.[15,16] A 2008 survey of 376 consultant dermatologists in Britain reported 49 deaths of patients taking methotrexate. Of these: one was caused by confusion between the 2.5 and 10mg doses, and two were caused by daily rather than weekly dosing.[17] However, this survey data relies upon recall over many years. One paper in 2006 reported 137 patient safety incidents related to methotrexate in England over the previous ten years.[18] However, these figures are likely to include a wide range of issues. National organisations in France, USA and Australia have also issued advice to prescribers, similar to the NHS; and, also similar to our findings, deaths and other errors associated with Methotrexate prescribing continue to occur in these countries.[19–21]

### Policy Implications and Interpretation

Previous work across several countries has resulted in a variety of suggested approaches to minimising the risks involved with methotrexate prescribing,[19,20,22] and the EMA is currently reviewing the issue.[4] NHS Improvement have classed overdoses associated with methotrexate as a Never Event; however, we can find no evidence of any action taken by any national NHS body when anomalous prescribing is detected in a region or practice. This is concerning, but may in part be explained by the fact that the responsibility for change is unclear. The “never event” framework was initially managed by the NPSA; this body closed in 2012 and became part of NHS Improvement, which does not have any remit over primary care; this may change when NHS Improvement moves to be part of NHS England in 2019.

Locally the NHS has invested extensively in ‘medicines optimisation’ activity, in which teams of pharmacists in every CCG monitor prescribing behaviour and advocate for change with individual clinicians. Previous work has shown that CCG membership is associated with prescribing patterns for practices [16,23,24] and in this study we found a significant relationship between the CCG to which a practice belongs and the variation in prescribing of methotrexate 10mg tablets. We are very concerned to see a number of CCGs exhibiting minimal change-related activity, in response to an important safety alert. In our view there is room for substantial improvement in personnel training for local staff, alongside open data monitoring by NHS England, and appropriate action for those failing to implement change.

Practices with a higher OpenPrescribing score were also more likely to have a higher proportion of 10mg tablets prescribed. It is unlikely that methotrexate causes poor prescribing in other areas or poor prescribing in other areas cause prescribing of methotrexate. We propose that both aspects of prescribing are linked by more fundamental issues, such as individual clinicians’ skills on evidence-based medicine; or the extent to which the practice team works together to review prescribing behaviour in their practice’s data, identify areas where they are outliers or exhibit unusual prescribing and take action collectively to address these issues.

### Future Research

The scale of breaches for methotrexate is clear. However it is likely that many other safety issues exhibit similarly prevalent breaches: given the low cost and high impact of data analyses to identify individual practices and the scale of national problems, we suggest this should be a high priority for research. While many deaths have been attributed to methotrexate, the data from death certificates is thin. Accessing and reviewing the text of coroners’ reports for all deaths associated with methotrexate would establish the role that methotrexate played in these deaths, and help identify preventive strategies: while this would take time, it seems consistent with the prioritisation of correct methotrexate dosing as a “Never Event”.

Decision support tools and “popups” at the point of care in a clinicians’ electronic health record software may offer an important opportunity to block unsafe prescribing. The methotrexate safety alert from the NPSA specifically highlights that EHRs should include “alerts and prompts”. The NHS makes significant investments in EHRs and it is imperative that their user interfaces support healthcare professionals to prescribe safely for patients. We found that one of EHR systems used in the NHS, Vision, was associated with higher prescribing of 10mg tablets, and this may reflect weaker preventive measures in this system. Our finding should be investigated immediately to understand if the design choice of the user interface in Vision is increasing the likelihood that a patient is prescribed 10mg tablets. More generally, we have been repeatedly blocked from researching the impact of “popups” on prescribing as there is no national framework or data on what popups are implemented in each setting. Additionally, this means that NHS commissioners and leaders cannot routinely identify what popups are implemented across the NHS. In order to realise the often cited potential of technology to improve safety the NHS [25,26] needs better oversight of technology and the ability to ensure system providers make modifications quickly if shortcomings are identified.

### Summary

The prevalence of unsafe methotrexate prescribing has gradually reduced, but it remains common, and with very substantial variation between GP practices. This is unlikely to be a unique problem. We recommend that the NHS invests in better strategies around audit and targeted dissemination of safety information, and identifies named individuals and roles with responsibility for implementing safety alerts.

#### Key Messages

##### What is already known about this subject?

- In the UK, it is recommended that when prescribing oral methotrexate tablets, only 2.5mg tablets should be used. This is to minimise the risk of accidental overdose, which can be fatal. Deaths and dosing errors associated with methotrexate prescribing continue to occur and the European Medicines Agency is currently reviewing the issue.

##### What does this study add?

- Breaches of this guidance are common, and vary widely between practices: 10% of all practices (697) give more than 14.3% of their methotrexate as 10mg tablets; and 1% of practices (66) give more than 52.4% as 10mg tablets.
- 21 deaths caused by methotrexate poisoning have been reported in England and Wales from 1993-2017.

##### How might this impact on clinical practice or future developments?

- The NHS should invest in better strategies around audit, targeted dissemination, and implementation of all safety information, not just for methotrexate. A full review of coroners reports for deaths associated with methotrexate poisoning should be conducted, to address any recurring themes and learning points
- Any interested party can view monthly data on all individual NHS GP practices breaching national methotrexate safety guidance at https://openprescribing.net/measure/methotrexate/

## Data Availability

Data are available in a public, open access repository on Github https://github.com/ebmdatalab/Methotrexate/releases/tag/v1.0 and Figshare https://figshare.com/collections/Methotrexate/4542308/1

## Acknowledgements

We are grateful to Peter Inglesby and Dave Evans for their contribution to maintaining databases and the OpenPrescribing website and Nicholas J. DeVito for his input on the analysis and paper draft.

## Conflicts of Interest

All authors have completed the ICMJE uniform disclosure form at http://www.icmje.org/conflicts-of-interest/ and declare the following: BG has received research funding from the Laura and John Arnold Foundation, the Wellcome Trust, the Oxford Biomedical Research Centre, the NHS National Institute for Health Research School of Primary Care Research, the Health Foundation and the World Health Organisation; he also receives personal income from speaking and writing for lay audiences on the misuse of science. RC, AJW, HC, SB are employed on BG’s grants for OpenPrescribing. BM is seconded to the DataLab from NHS England.

## Contributorship

BG conceived the study. BG BM HC AW RC SB designed the methods. BM collected and analysed the data with methodological and interpretation input from BG AW HC SB RC. BM drafted the manuscript. All authors contributed to and approved the final manuscript. BG supervised the project and is guarantor.

## Funding

No specific funding was obtained for this project. Funders had no role in the study design, collection, analysis, and interpretation of data; in the writing of the report; and in the decision to submit the article for publication.

## Ethical approval

This study uses open, publicly available data, and data publically available on request under the Freedom of Information Act therefore no ethical approval was required.

## Guarantor

BG is guarantor.

## Appendix

### Measures included in composite prescribing score

- Antibiotic stewardship: co-amoxiclav, cephalosporins & quinolones (KTT9),
- Antibiotic stewardship: three-day courses for uncomplicated UTIs (KTT9),
- Antibiotic stewardship: volume of antibiotic prescribing (KTT9),
- Ciclosporin and tacrolimus oral preparations prescribed generically,
- Co-proxamol,
- Desogestrel prescribed as a branded product,
- Diltiazem preparations (>60mg) prescribed generically,
- Extended-release quetiapine,
- Glaucoma eye drops prescribed by brand’,
- High-cost ACE inhibitors,
- High-cost ARBs,
- High-cost drugs for erectile dysfunction,
- High-cost PPIs,
- High-cost statins,
- High dose inhaled corticosteroids,
- High dose opioids as percentage regular opioids,
- High dose opioids per 1000 patients,
- Higher dose Proton Pump Inhibitors (PPIs),
- Keppra vs. levetiracetam,
- Long-acting insulin analogues (KTT12),
- Low and medium intensity statins,
- Methotrexate 10 mg tablets,
- Nebivolol 2.5mg tablets,
- NHS England Low Priority Treatment - All Low Priority Treatments,
- Non-preferred NSAIDs and COX-2 inhibitors (KTT13),
- Other lipid-modifying drugs,
- Pregabalin prescribed as Lyrica,
- Prescribing of dipyridamole,
- Prescribing of high cost tramadol preparations,
- Prescribing of trimethoprim vs nitrofurantoin,
- Short acting beta agonist inhalers,
- Silver dressings,
- Soluble/effervescent forms of paracetamol and co-codamol,
- Topical treatment of fungal nail infections,
- Vitamin B complex,
- Anxiolytics and Hypnotics: Average Daily Quantity per 1000 patients,
- Anxiolytics and Hypnotics: Average Daily Quantity per item,
- Prescribing of opioids (total oral morphine equivalence)

These measures can be viewed online at https://openprescribing.net/measure/

## Notes

### Author Declarations

All relevant ethical guidelines have been followed and any necessary IRB and/or ethics committee approvals have been obtained.

Any clinical trials involved have been registered with an ICMJE-approved registry such as ClinicalTrials.gov and the trial ID is included in the manuscript.

## References

1 Joint Formulary Committee. British National Formulary. https://bnf.nice.org.uk/ (accessed 13 Jul 2018).

2 Cambridgeshire Health Authority. Methotrexate Toxicity:An Inquiry into the death of a Cambridgeshire patient. http://anthonycox.org/wp-content/uploads/2012/12/methotrexate-toxicity.pdf (accessed 25 Jul 2018).

3 National Patient Safety Agency. Alert – Improving compliance with oral methotrexate guidelines 2006. Specialist Pharmacy Service. 2006.https://www.sps.nhs.uk/articles/npsa-alert-improving-compliance-with-oral-methotrexate-guidelines-2006/ (accessed 25 Jul 2018).

4 European Medicines Agency. EMA reviewing risk of dosing errors with methotrexate. 2018. https://www.ema.europa.eu/documents/press-release/ema-reviewing-risk-dosing-errors-methotrexate_en.pdf (accessed 14 Jan 2019).

5 NHS Improvement. Revised Never Events policy and framework. https://improvement.nhs.uk/resources/never-events-policy-and-framework/ (accessed 14 Jan 2018).

6 NHS Improvement. Recommendations from NPSA alerts that remain relevant to Never Events. https://improvement.nhs.uk/documents/2267/Recommendations_from_NPSA_alerts_that_remain_relevant_to_NEs_FINAL.pdf

7 EBM Datalab. Methotrexate 10 mg tablets by all CCGs - OpenPrescribing. OpenPrescribing.net. 2018. https://openprescribing.net/measure/methotrexate/ (accessed 14 Jan 2019).

8 NHS Business Services Authority. Information Services Portal (ISP). NHS Business Services Authority. https://www.nhsbsa.nhs.uk/information-services-portal-isp (accessed 25 Jul 2018).

9 NHS Digital. Patients registered at a GP practice. https://digital.nhs.uk/data-and-information/data-tools-and-services/data-services/general-practice-data-hub/patients-registered-at-a-gp-practice (accessed 14 Jan 2019).

10 NHS Business Services Authority. Freedom of Information Request: 7951. https://apps.nhsbsa.nhs.uk/FOI/foiRequestDetail.do?bo_id=7951 (accessed 28 Jan 2019).

11 Office for National Statistics. Freedom of Information Request:Number of deaths relating to methotrexate, England and Wales, deaths registered between 1993 and 2017. 2018.https://www.ons.gov.uk/peoplepopulationandcommunity/birthsdeathsandmarriages/deaths/adhocs/008937numberofdeathsrelatingtomethotrexateenglandandwalesdeathsregisteredbetween1993and2017 (accessed 28 Jan 2019).

12 Walker AJ, Croker R, Bacon S, et al. Is use of homeopathy associated with poor prescribing in English primary care? A cross-sectional study. J R Soc Med 2018;111:167–74. doi:10.1177/0141076818765779

13 Methotrexate. EBM Datalab - GitHub Repository. 2019.https://github.com/ebmdatalab/Methotrexate/releases/tag/v1.0

14 MacKenna B. Methotrexate - FOI Repository. 2019. doi:10.6084/M9.FIGSHARE.C.4542308.V1

15 Walker AJ, Bacon S, Curtis H, et al. Six months on: NHS England needs to focus on dissemination, implementation and audit of its low-priority initiative. J R Soc Med 2018;:141076818808429. doi:10.1177/0141076818808429

16 Croker R, Walker AJ, Goldacre B. Why did some practices not implement new antibiotic prescribing guidelines on urinary tract infection? A cohort study and survey in NHS England primary care. J Antimicrob Chemother Published Online First: 22 December 2018. doi:10.1093/jac/dky509

17 Collin B, Srinathan SK, Finch TM. Methotrexate: prescribing and monitoring practices among the consultant membership of the British Association of Dermatologists. Br J Dermatol 2008;158:793–800. doi:10.1111/j.1365-2133.2007.08426.x

18 Grills C, Burge S. Methotrexate: improving safety profile. Australas J Dermatol 2006;47:178–81. doi:10.1111/j.1440-0960.2006.00267.x

19 Grissinger M. Severe Harm and Death Associated With Errors and Drug Interactions Involving Low-Dose Methotrexate. P T 2018;43:191–248.https://www.ncbi.nlm.nih.gov/pubmed/29622936

20 Cairns R, Brown JA, Lynch A-M, et al. A decade of Australian methotrexate dosing errors. Med J Aust 2016;204:384.https://www.ncbi.nlm.nih.gov/pubmed/27256650

21 Vial T, Patat AM, Boels D, et al. Adverse consequences of low-dose methotrexate medication errors: data from French poison control and pharmacovigilance centers. Joint Bone Spine Published Online First: 19 September 2018. doi:10.1016/j.jbspin.2018.09.006

22 Goldsmith P, Roach A. Methods to enhance the safety of methotrexate prescribing. J Clin Pharm Ther 2007;32:327–31. doi:10.1111/j.1365-2710.2007.00834.x

23 Walker AJ, Curtis HJ, Bacon S, et al. Trends, geographical variation and factors associated with prescribing of gluten-free foods in English primary care: a cross-sectional study. BMJ Open 2018;8:e021312. doi:10.1136/bmjopen-2017-021312

24 Walker AJ, Curtis HJ, Bacon S, et al. Trends and variation in prescribing of low-priority treatments identified by NHS England: a cross-sectional study and interactive data tool in English primary care. J R Soc Med 2018;111:203–13. doi:10.1177/0141076818769408

25 Donaldson LJ, Appleby L, Boyce J. An organisation with a memory: report of an expert group on learning from adverse events in the NHS. 2000.https://www.aagbi.org/sites/default/files/An%20organisation%20with%20a%20memory.pdf

26 The future of healthcare: our vision for digital, data and technology in health and care. GOV.UK. 2018.https://www.gov.uk/government/publications/the-future-of-healthcare-our-vision-for-digital-data-and-technology-in-health-and-care/the-future-of-healthcare-our-vision-for-digital-data-and-technology-in-health-and-care (accessed 13 Jan 2019).

